# Comprehensive Spatiotemporal Analysis of Opioid Poisoning Mortality in Ohio from 2010 to 2016

**DOI:** 10.1101/19005454

**Authors:** Chihyun Park, Jean R. Clemenceau, Anna Seballos, Sara Crawford, Rocio Lopez, Tyler Coy, Gowtham Atluri, Tae Hyun Hwang

**Author notes:** Corresponding authors: Gowtham Atluri, Dept. of EECS, University of Cincinnati, P.O. Box 210030, Cincinnati, OH 45221, USA, Tae Hyun Hwang, PhD, Dept. of QHS, Lerner Research Institute, Cleveland Clinic, 9500 Euclid Ave, Cleveland, OH 44195, USA. these authors contributed equally to this work (Chihyun Park, Jean R. Clemenceau).

## Abstract

**Objective:** We aimed to identify (1) differences in opioid poisoning mortality among population groups, (2) geographic clusters of opioid-related deaths over time, and (3) health conditions co-occurring with opioid-related death in Ohio by computational analysis.

**Materials and Methods:** We used a large-scale Ohio vital statistic dataset from the Ohio Department of Health (ODH) and U.S. Census data from 2010-2016. We surveyed population differences with demographic profiling and use of relative proportions, conducted spatiotemporal pattern analysis with spatial autocorrelation via Moran statistics at the census tract level, and performed comorbidity analysis using frequent itemset mining and association rule mining.

**Results:** Our analyses found higher rates of opioid-related death in people aged 25-54, whites, and males. We also found that opioid-related deaths in Ohio became more spatially concentrated during 2010-2016, and tended to be most clustered around Cleveland, Columbus and Cincinnati. Drug abuse, anxiety and cardiovascular disease were found to predict opioid-related death.

**Discussion:** Comprehensive data-driven spatiotemporal analysis of opioid-related deaths provides essential identification of demographic, geographic and health factors related to opioid abuse. Future research should access personal health information for more detailed comorbidity analysis, as well as expand spatiotemporal models for real-time use.

**Conclusion:** Computational analyses revealed demographic differences in opioid poisoning, changing regional patterns of opioid-related deaths, and health conditions co-occurring with opioid overdose for Ohio from 2010-2016, providing essential knowledge for both government officials and caregivers to establish policies and strategies to best combat the opioid epidemic.

## INTRODUCTION

Opioids are a class of drugs derived from the opium poppy plant that block the reception of pain signals in the brain. Opioids include prescription pain relievers such as oxycodone, hydrocodone, codeine, and morphine and are used to alleviate chronic pain and manage postoperative pain. Opioids are also used illegally, including heroin and synthetic opioids such as fentanyl (which is also used medically). While the number of opioid prescriptions written has skyrocketed in the last two decades, illegal opioid use, opioid abuse and accidental opioid overdose have also increased.[1] Opioid-related overdoses are commonly referred to and categorized as “opioid poisoning (T40.0-T40.4, T40.6)” according to International Classification of Disease (ICD) codes (ICD-10).[2] The mortality rate due to opioid poisoning has consistently increased each year and now it is recognized as a national crisis.[3] Since 2000, death from drug overdoses has increased by 137%, while deaths from opioid overdoses has increased by 200%.[4] According to the data released by the U.S Department of Health and Human Services, 130 people died each day from opioid-related drug overdoses in 2017 and 11.5 million people misused prescription opioids.[5]

The opioid epidemic is a nationwide issue, and scientific research is key to identifying causal factors, discovering susceptible communities and developing policies to address this problem. Increased availability of healthcare data presents a tremendous opportunity for data analysis to identify potential predictors of opioid misuse and predict patients susceptible to opioid misuse.[6] There have been several attempts to apply computational or statistical analysis to solve the problems associated with the opioid epidemic.[7-11] The purposes of these studies varied, ranging from understanding and detecting geographical clusters or hot spots where opioid misuse has occurred within urban neighborhoods,[12] to developing a surveillance model to identify patients who are misusing opioids or are over-prescribed.[10] Spatiotemporal analysis has been applied to identify relationships between environmental or economic factors and opioid misuse or overdose.[12-14] Through these attempts, several predictive covariates were revealed. Housing vacancy, dilapidated housing and misdemeanor arrests have been shown to be significantly associated with illicit drug activity.[12, 13] Additionally, individuals with a higher income and greater access to healthcare have been found to be associated with prescription opioid poisoning.[14] Existing research on comorbidity with opioid abuse mostly focuses on psychiatric disorders and co-occurring substance abuse.[7-9,15-18] Logistic regression models have been developed to predict patients who may be susceptible to prescription opioid abuse.[7-9] Comorbid psychiatric disorders have been considered for their impact on the efficacy of various treatments of opioid use disorder,[15] and literature reviews have found high rates of co-occurrence between opioid use disorder and anxiety, psychiatric comorbidity, and drug use.[16-18]

These studies focus on either city-level spatiotemporal patterns, and/or solely on abuse of a single class of opioids. Our research performs comprehensive profiling, spatial pattern mining, and comorbidity analysis on a large-sized dataset collected at a state-wide level for all opioid-related deaths. Our research utilizes a large-scale vital statistic dataset from the Ohio Department of Health (ODH) and U.S. Census datasets to perform retrospective and extensive analysis at a statewide and census tract level. The purpose of this study is to identify demographic and spatial factors, as well as co-occurring health conditions that are associated with opioid-related overdose. To achieve this, we applied several data mining approaches such as clustering and frequent pattern mining. We expect that our results will inform policy makers, law enforcement, emergency health services, and caregivers to predict and prevent opioid abuse throughout Ohio.

## METHODS

### Data description

In this paper, we used ODH mortality data containing decedent information by opioid poisoning and misuse. We were approved to access limited and identifiable data, completed the Data Use Agreement (DUA), and de-identified decedent data. Access and use of the data was approved by the ODH IRB. We filtered ODH mortality data by selecting only opioid-related records. We considered a death opioid-related if the record had a positive value for any of the following nine indicators: Methadone, opiates, prescription opiates, Fentanyl, Fentanyl and Analogues, Carfentanil, designer opioids, commonly prescribed opioids, and/or other opioids. Then, we chose records for analysis which contained positive values for “Opioid Related Death,” “Unintended Death,” “Undetermined death,” and “Drug Poisoning Injury Mechanism.”

As a result, 13,094 records satisfied these conditions from 2010-2016. Of these records, 13,057 were not missing any values in the demographic and location fields. Figure 1 shows the entire process of data refinement. This dataset was used in our comorbidity analysis mentioned in the next section. This dataset was used for the remaining analysis and is hereafter referred to as “ODH-Opioid data.” We obtained publicly available demographic data from the 2010 U.S. Census Bureau and from the American Community Survey (ACS) from 2011-2016 to gather characteristics of the entire population of Ohio, as well as from each Ohio census tract.[19]

**Figure 1.**
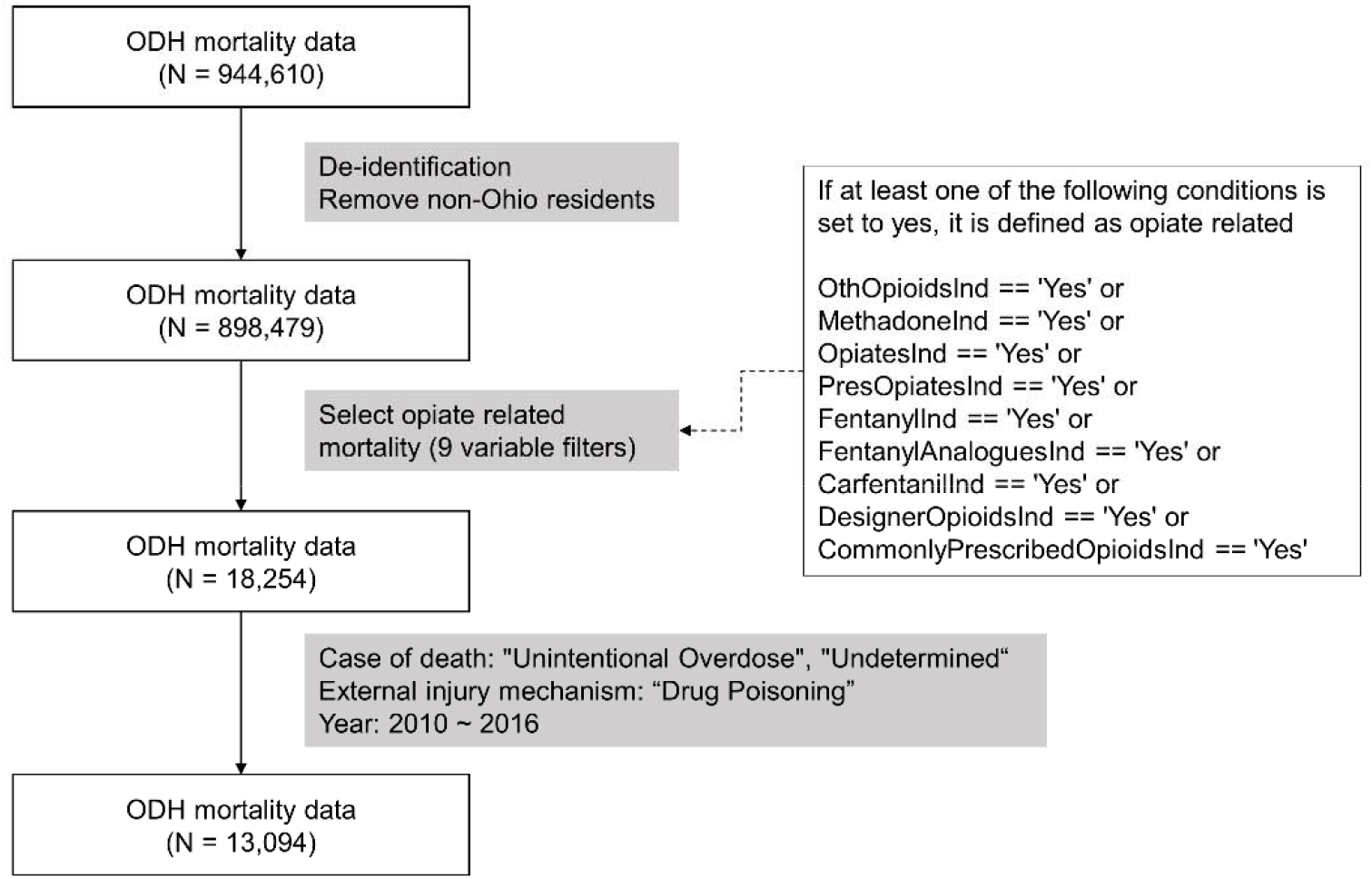
The process of data refinement. From the entire ODH mortality data, we finally extracted 13,094 records related with opioid poisoning.

### Analysis

Our analysis of 2010-2016 opioid overdose data is composed of the following three sections: 1) demographic-based analysis to survey differences among population groups; 2) spatiotemporal pattern analysis (census tract level) to discover potential global and local spatial clusters; 3) co-occurrence analysis with cause of death and its corresponding health condition to identify potential patterns or rules between them. As the data covers a seven-year period, it is possible to confirm trends and patterns over the years.

1. Demographic-based analysis to survey disparities among population groups

Decedent demographic information, such as age, race and sex, was taken from ODH-Opioid data. Then, the same population-level demographic data in Ohio was obtained from the ACS. Based on these datasets, we compared and analyzed the prevalence of opioid-induced deaths in population groups relative to population distribution. We calculated relative proportions for each population group based on these two datasets for seven years. Then, we performed a paired t-test to investigate the statistical difference between the proportion values of two datasets.

2.Spatiotemporal pattern analysis (census tract level) to discover potential global and local spatial clusters

Each record in ODH-Opioid data contains geographical information about a given decedent, such as residence or place of death. It is necessary to investigate whether deaths tend to be focused in a specific region or dispersed within a dataset to predict spatial patterns of opioid abuse. To achieve this, we applied spatial autocorrelation, which can measure the degree to which an object is similar to other nearby objects. More specifically, a positive value of spatial autocorrelation indicates the relationship between the value at a location and the values of its neighbors is positive; otherwise, the spatial autocorrelation is negative. Moran statistics are one of the commonly used approaches to measure this relationship.[20] There are two types of Moran’s *I* statistics: global and local.

Global Moran’s *I* statistics provide a single measure of spatial autocorrelation for an attribute in a region as a whole, whereas Local Moran’s *I* statistics provide a measure of the tendency of a given unit to have an attribute value that is correlated with values of nearby areas. Local spatial autocorrelation analysis is based on LISA (Local Indicators of Spatial Autocorrelation) statistics and is computed for each individual unit. The following two functions represent Global and Local Moran’s *I* statistics:

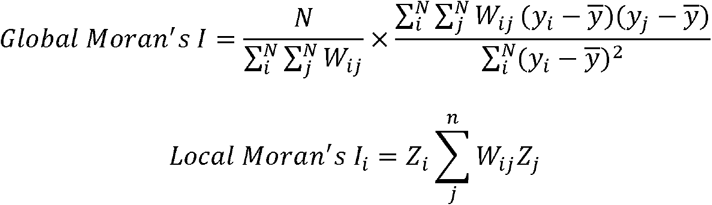

*N* indicates the number of units in the whole region, *y* is the attribute value for each unit, *W* indicates weight matrix (*W*ij represent connectivity between unit *i* and *j*) and Z is the standardized score of attribute values for the corresponding unit. In Local Moran’s *I, j* indicates neighboring units of *i*. Informally, +1 indicates strong positive spatial autocorrelation (i.e., clustering of similar values), 0 indicates random spatial ordering and −1 indicates strong negative spatial autocorrelation (perfect dispersion, i.e., a checkerboard pattern).

In our analysis, we used census tract as the unit of region. Census tracts are small, relatively permanent statistical subdivisions within a county. We aggregated the number of deaths by census tract level and then normalized the number of deaths by dividing the total population in the corresponding Tract ID. This normalization allowed spatial autocorrelation to be performed with the ratio of the actual population to the number of deaths in the area. We used these census tract death rates for spatiotemporal pattern analysis.

We performed spatiotemporal pattern analysis by using a free, open source software tool, GeoDa.[21] We applied Global Moran’s *I* statistics to Ohio at the state-level and LISA statistics to each region by census tract. GeoDa also reported p-values to show statistical significance, and we applied 0.05 as our alpha level.

3.Comorbidity analysis with cause of death and its corresponding health condition

This analysis aimed to identify which health conditions frequently co-occurred with opioid poisoning, and whether the cause of death exhibited significant associations with co-occurring health conditions. We applied frequent itemset mining and association rule mining to ODH-Opioid data to carry out these analyses. Although ODH-Opioid data are filtered as “death by opioid misuse,” the specific causes of death are more diverse. In the ODH mortality dataset, cause of death was listed by ICD codes (ICD-9 and ICD-10). In order to analyze the observed health conditions, we had to simplify the literal text as reported for the deceased person to a standard term. Figure 2 shows the overall workflow for the text clustering method we used to convert the literal text to a structured term. Then, we integrated cause of death with converted health condition and applied frequent itemset mining to reveal which events (i.e. itemset) frequently occur together.[22]

**Figure 2.**
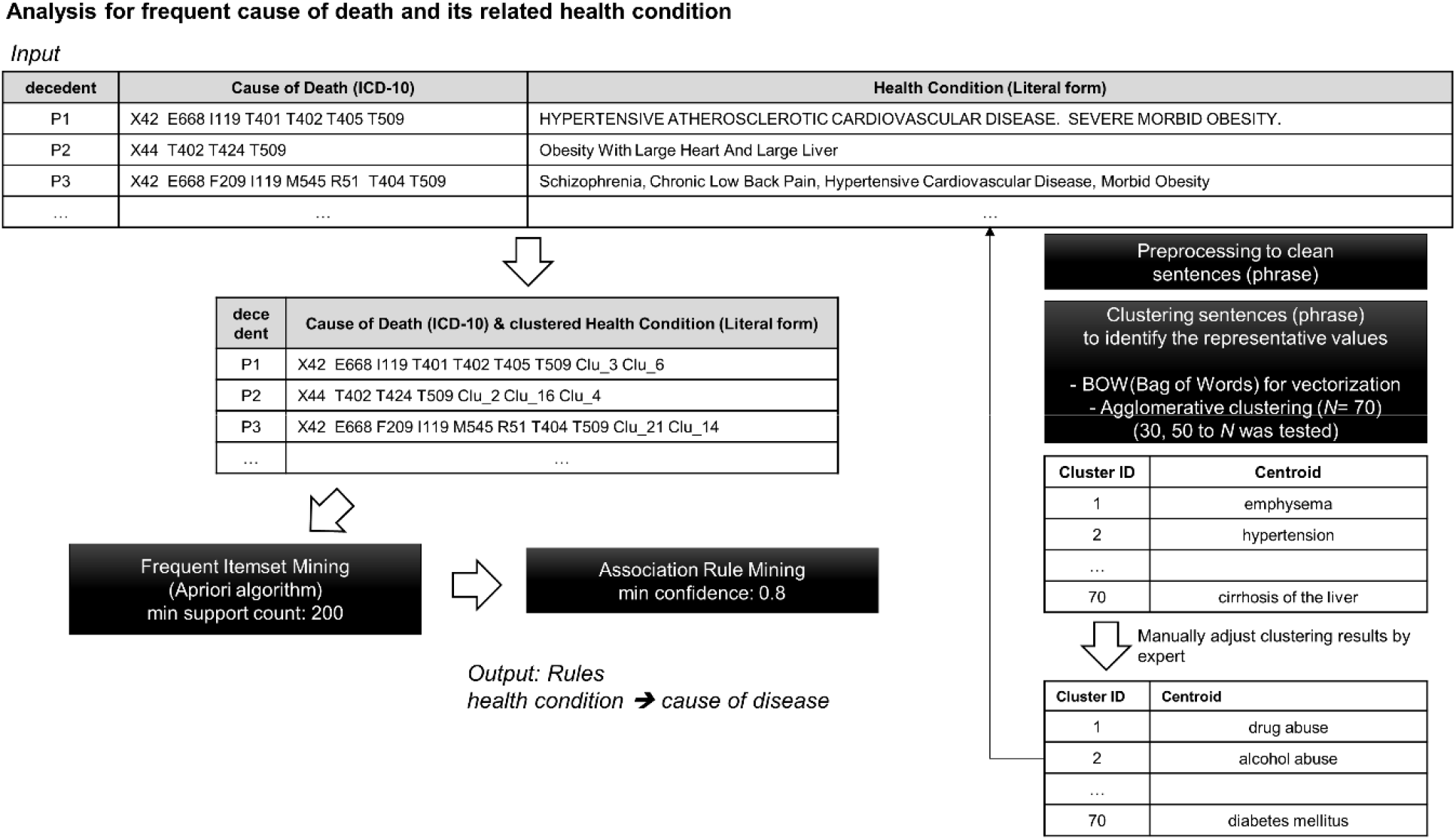
Entire workflow of co-occurrence analysis with the list of cause of death and corresponding health condition.

From the results of our frequent itemset mining, we performed association rule mining with a minimum confidence of 0.5 in order to quantify the association between two itemsets. After sorting all rules by Lift value, only the rules with at least one health condition in itemset A and at least one cause of death in itemset B were selected from the results (Lift > 1: positive relationship, which means if A is present, then B happens).

## RESULTS

### 1. Demographic-based analysis to survey disparities among population groups

Using ODH-Opioid data, we compared the demographics between the overall Ohio population and those who died due opioid poisoning. Figure 3 (A) shows the difference between population proportion and mortality proportion by opioid poisoning across seven years for each age group. For example, while the group aged 24-44 years made up an average of 25.1% of the overall population across the 7 years, the same age group consisted of an average of 51.3% of the opioid deaths. There was a significant difference between the two groups for all ages except the 55-60 age group. This result implies that the pattern of mortality by opioid poisoning is different from the general population distribution. In particular, groups aged 25-44 and 45-54 years died by opioid poisoning more frequently than any other age groups compared to the general population distribution. We performed the same analysis for race and sex. Figures 3 (B) and (C) show the comparison results. There was a statistically significant difference identified between the mortality and population distribution for all races except for “Native Hawaiian or Other Pacific Islander,” with the white population exhibiting the highest mortality counts. Similarly, there was a statistically significant difference for both males and females, with a higher fatality proportion for males, and a lower fatality proportion for females, as compared to their respective population proportions. In summary, between 2010-2016 in Ohio, people who were white, male and aged 25-54 years were most likely to die by opioid poisoning. Table 1 shows detailed results of the statistical tests.

**Table 1.**
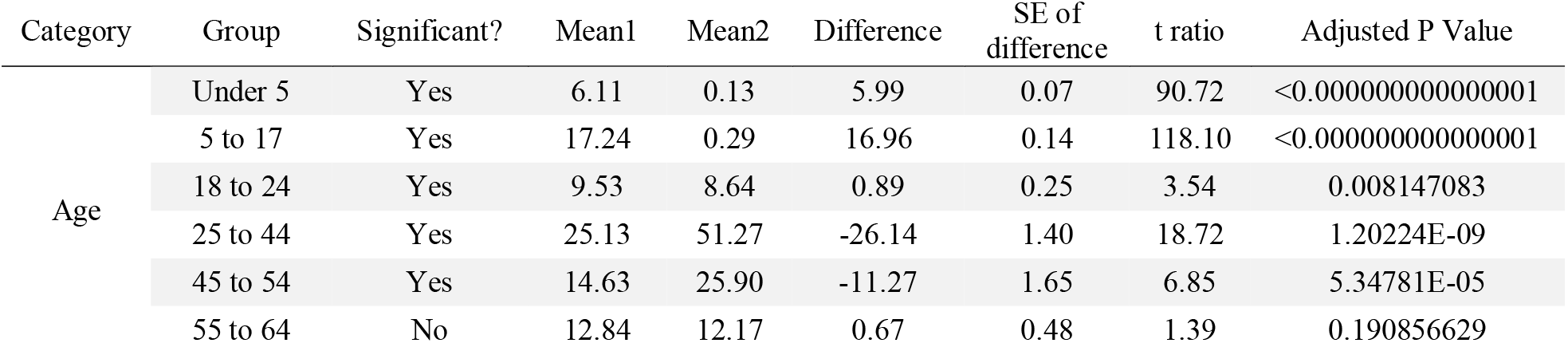
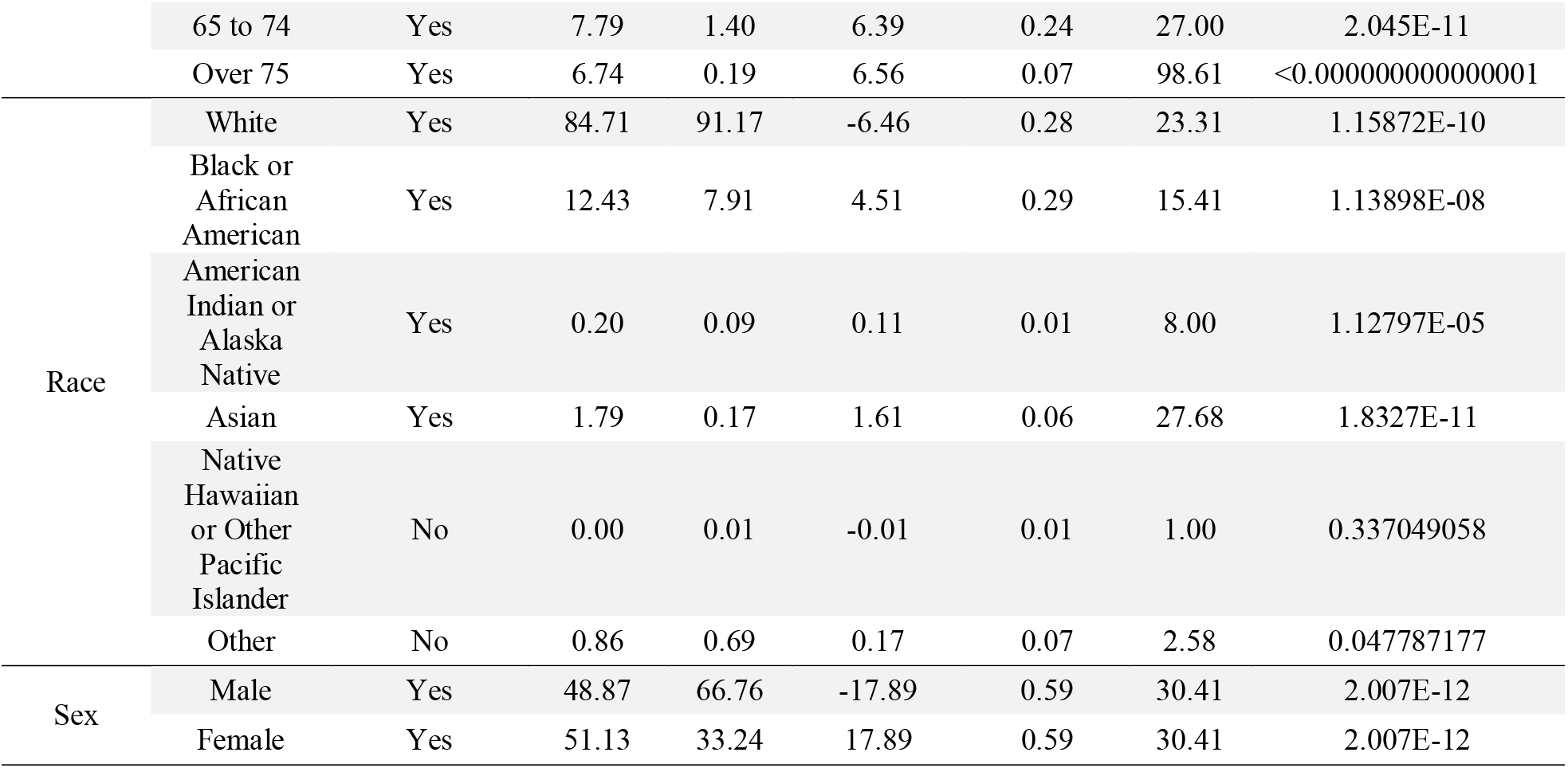
Detailed results of the statistical tests for investigating disparities between population groups.

| Category | Group | Significant? | Mean1 | Mean2 | Difference | SE of difference | t ratio | Adjusted P Value |
| --- | --- | --- | --- | --- | --- | --- | --- | --- |
| Age | Under 5 | Yes | 6.11 | 0.13 | 5.99 | 0.07 | 90.72 | <0.0000000000000001 |
|  | 5 to 17 | Yes | 17.24 | 0.29 | 16.96 | 0.14 | 118.10 | <0.0000000000000001 |
|  | 18 to 24 | Yes | 9.53 | 8.64 | 0.89 | 0.25 | 3.54 | 0.008147083 |
|  | 25 to 44 | Yes | 25.13 | 51.27 | -26.14 | 1.40 | 18.72 | 1.20224E-09 |
|  | 45 to 54 | Yes | 14.63 | 25.90 | -11.27 | 1.65 | 6.85 | 5.34781E-05 |
|  | 55 to 64 | No | 12.84 | 12.17 | 0.67 | 0.48 | 1.39 | 0.190856629 |
|  | 65 to 74 | Yes | 7.79 | 1.40 | 6.39 | 0.24 | 27.00 | 2.045E-11 |
|  | Over 75 | Yes | 6.74 | 0.19 | 6.56 | 0.07 | 98.61 | <0.000000000000001 |
| Race | White | Yes | 84.71 | 91.17 | -6.46 | 0.28 | 23.31 | 1.15872E-10 |
|  | Black or African American | Yes | 12.43 | 7.91 | 4.51 | 0.29 | 15.41 | 1.13898E-08 |
|  | American Indian or Alaska Native | Yes | 0.20 | 0.09 | 0.11 | 0.01 | 8.00 | 1.12797E-05 |
|  | Asian | Yes | 1.79 | 0.17 | 1.61 | 0.06 | 27.68 | 1.8327E-11 |
|  | Native Hawaiian or Other Pacific Islander | No | 0.00 | 0.01 | -0.01 | 0.01 | 1.00 | 0.337049058 |
|  | Other | No | 0.86 | 0.69 | 0.17 | 0.07 | 2.58 | 0.047787177 |
| Sex | Male | Yes | 48.87 | 66.76 | -17.89 | 0.59 | 30.41 | 2.007E-12 |
|  | Female | Yes | 51.13 | 33.24 | 17.89 | 0.59 | 30.41 | 2.007E-12 |

**Figure 3.**
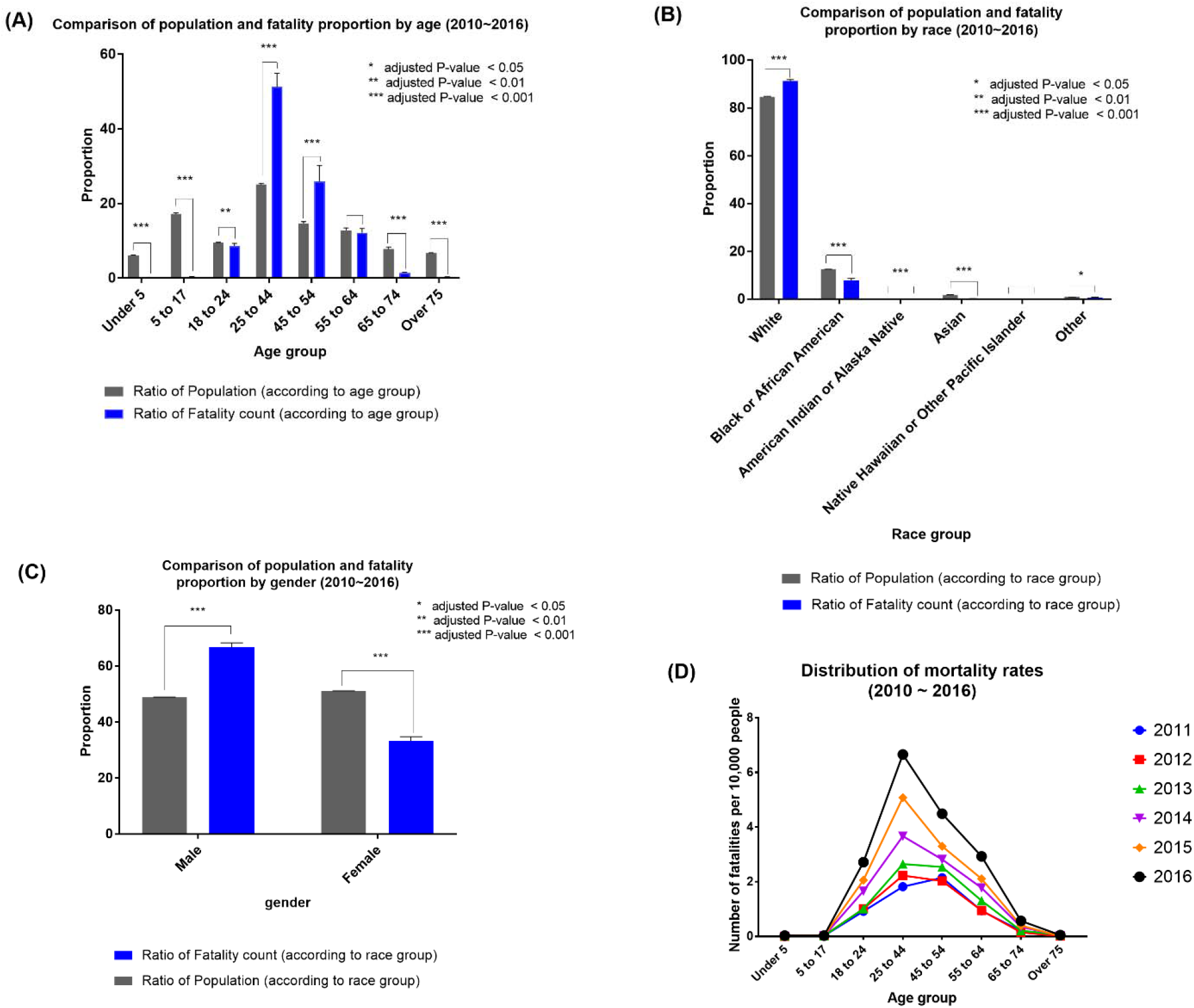
Comparison of population and mortality proportion by several demographic factors.

Finally, as shown in Figure 3 (D), we calculated the mortality rate per 10,000 people for each age group in each year. As we concluded earlier, morality rates for age groups 25-44 and 45-54 were the highest and the ratio increased with time.

### 2. Spatiotemporal pattern analysis (census tract level) to discover potential global and local spatial clusters

As seen in Figure 4, the Global Moran’s I value, which provides a single measure of spatial autocorrelation for an attribute in a region as a whole, continuously increased from 2010-2016. This signifies higher specificity and lower randomization of clustering, meaning that the opioid crisis became more spatially concentrated during these years.

**Figure 4.**
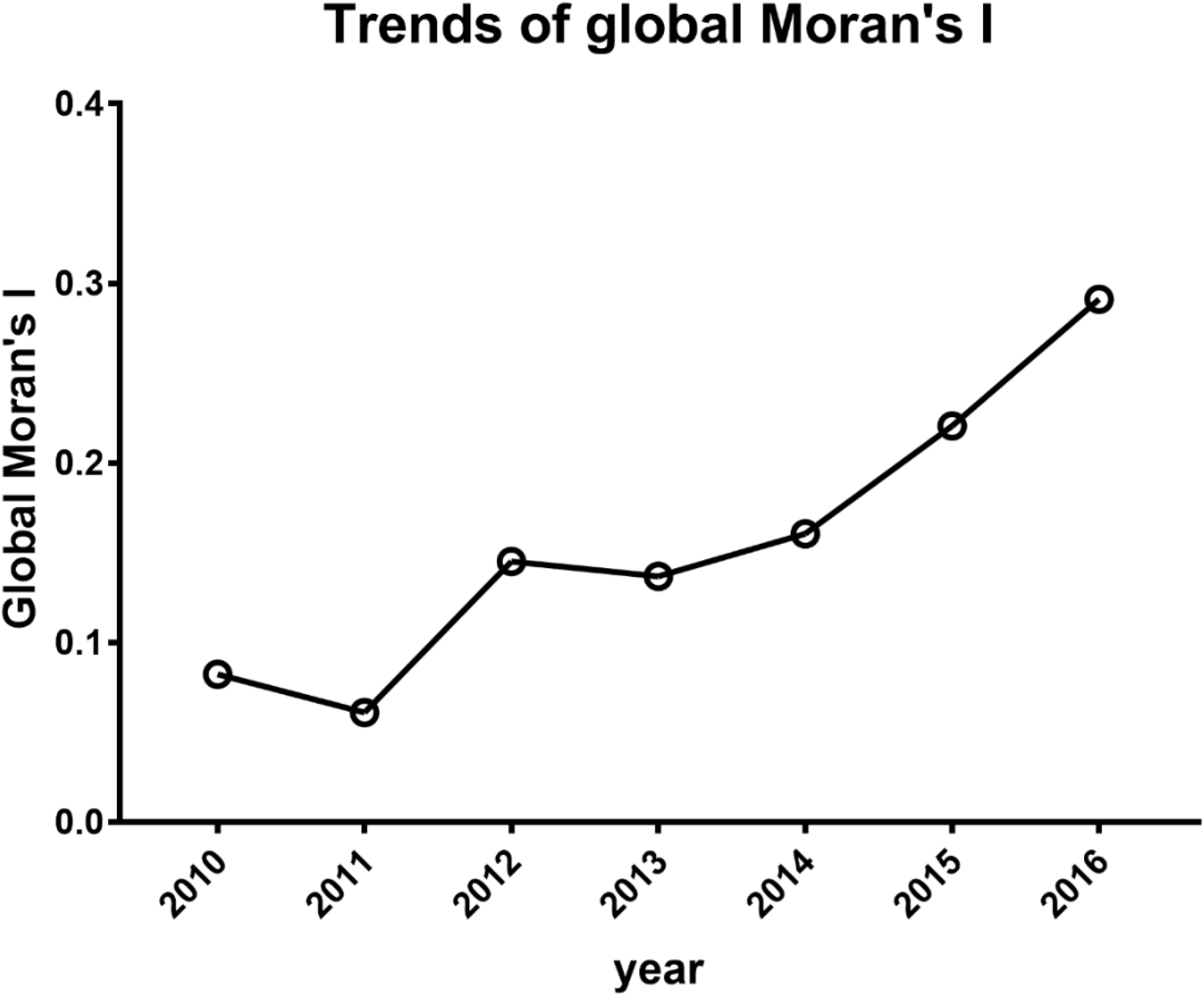
Changing pattern of Global Moran’s I statistics for seven years in Ohio, 2010-2016.

Figure 5 demonstrates the results of our spatiotemporal pattern analysis for each census tract. As shown in Figure 5, there are four types of patterns of LISA statistics as follows: High-High, High-Low, Low-High, and Low-Low. The first field of a given LISA pattern indicates the degree of LISA statistics in the corresponding region and the second field of a given LISA pattern indicates the degree of LISA statistics of the neighboring regions. For example, a High-Low designation for a certain region means the LISA statistics of the region is significantly high but the values of its neighboring regions are significantly low. Generally, we can consider High-High and Low-Low status as spatial clusters because these regions have similar LISA statistics values. It was revealed that the High-High and Low-Low regions have become increasingly dispersed from 2010 to 2016.

**Figure 5.**
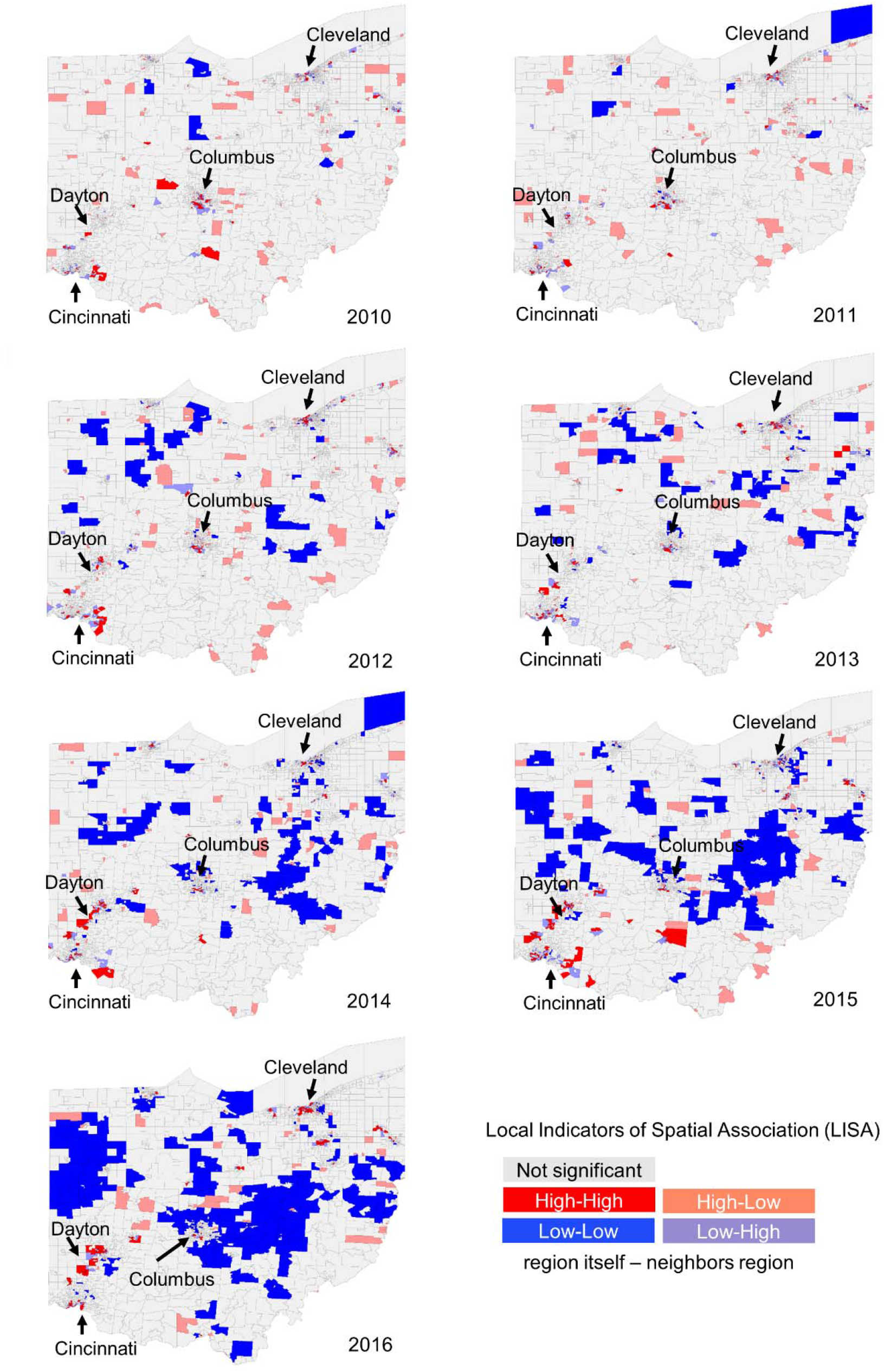
Spatial clusters with normalized mortality (number of deaths divided by number of population) for opioid poisoning by census tract, Ohio, 2010-2016.

Table 2 shows the top 10 census tracts represented by county name based on the LISA statistics value. As a result, it was confirmed that the counties located inside or near Cleveland, Columbus, and Cincinnati, which are the most populated in Ohio, had the highest concentration of deaths by opioid poisoning.

**Table 2.** List of top 10 census tracts and their corresponding county by statistics value from 2010 to 2016. Three large cities, Cleveland, Columbus and Cincinnati, experienced more clustering of opioid poisoning than any other regions.

| Year | 2010 | 2011 | 2012 | 2013 | 2014 | 2015 | 2016 |
| --- | --- | --- | --- | --- | --- | --- | --- |
| Representing<br>with County<br>for top 10<br>Tract by<br>LISA | Hamilton | Hamilton | Hamilton | Cuyahoga | Butler | Butler | Summit |
|  | Franklin | Hamilton | Cuyahoga | Butler | Butler | Butler | Montgomery |
|  | Franklin | Cuyahoga | Montgomery | Butler | Butler | Hamilton | Cuyahoga |
|  | Cuyahoga | Hamilton | Franklin | Cuyahoga | Hamilton | Butler | Montgomery |
|  | Franklin | Hamilton | Montgomery | Cuyahoga | Butler | Clark | Summit |
|  | Ross | Franklin | Cuyahoga | Butler | Butler | Montgomery | Summit |
|  | Franklin | Franklin | Cuyahoga | Cuyahoga | Hamilton | Hamilton | Montgomery |
|  | Franklin | Cuyahoga | Hamilton | Cuyahoga | Butler | Clark | Summit |
|  | Franklin | Clark | Cuyahoga | Cuyahoga | Butler | Butler | Summit |
|  | Franklin | Cuyahoga | Mahoning | Cuyahoga | Butler | Butler | Butler |
| Most<br>Popular<br>County | Franklin<br>County:<br>7/10<br>(in<br>Columbus) | Hamilton<br>County:<br>4/10<br>(in<br>Cincinnati) | Cuyahoga<br>County:<br>4/10<br>(in<br>Cleveland) | Cuyahoga<br>County:<br>7/10<br>(in<br>Cleveland) | Butler<br>County:<br>8/10<br>(in<br>Cincinnati) | Butler<br>County:<br>5/10<br>(in<br>Cincinnati) | Summit<br>County:<br>5/10<br>(in Akron) |
| LISA_I<br>(P-value <<br>0.05) | 22.713 | 10.792 | 15.508 | 15.683 | 33.174 | 26.137 | 19.071 |
|  | 7.835 | 6.930 | 15.091 | 13.258 | 28.665 | 17.406 | 18.977 |
|  | 5.897 | 6.305 | 15.000 | 12.516 | 13.559 | 17.028 | 14.672 |
|  | 5.316 | 5.288 | 13.030 | 11.431 | 11.540 | 16.645 | 13.514 |
|  | 5.052 | 5.068 | 11.734 | 10.880 | 11.525 | 13.854 | 12.387 |
|  | 4.521 | 4.610 | 11.374 | 10.306 | 11.471 | 13.344 | 11.337 |
|  | 4.203 | 4.243 | 10.623 | 7.317 | 11.382 | 11.492 | 11.096 |
|  | 3.846 | 3.745 | 9.057 | 7.185 | 11.199 | 9.197 | 10.254 |
|  | 3.716 | 3.654 | 8.903 | 7.074 | 6.632 | 8.781 | 9.744 |
|  | 3.447 | 3.499 | 8.203 | 6.606 | 6.565 | 8.055 | 9.102 |

### 3. Co-occurrence analysis with cause of death and its corresponding health condition

Table 3 shows the representative terms for health conditions of the deceased derived from a text clustering approach. As mentioned in the Methods section, we used these health condition terms and cause of death classified by ICD-10 code as an input of frequent itemset and pattern mining to reveal the underlying relationship between health condition and cause of death with regard to opioid poisoning mortality. Of the total 13,094 records, only 2,887 records contained data for both health condition and cause of death, so the remaining records which had a value of “NA” were excluded for the co-occurrence analysis.

**Table 3.**
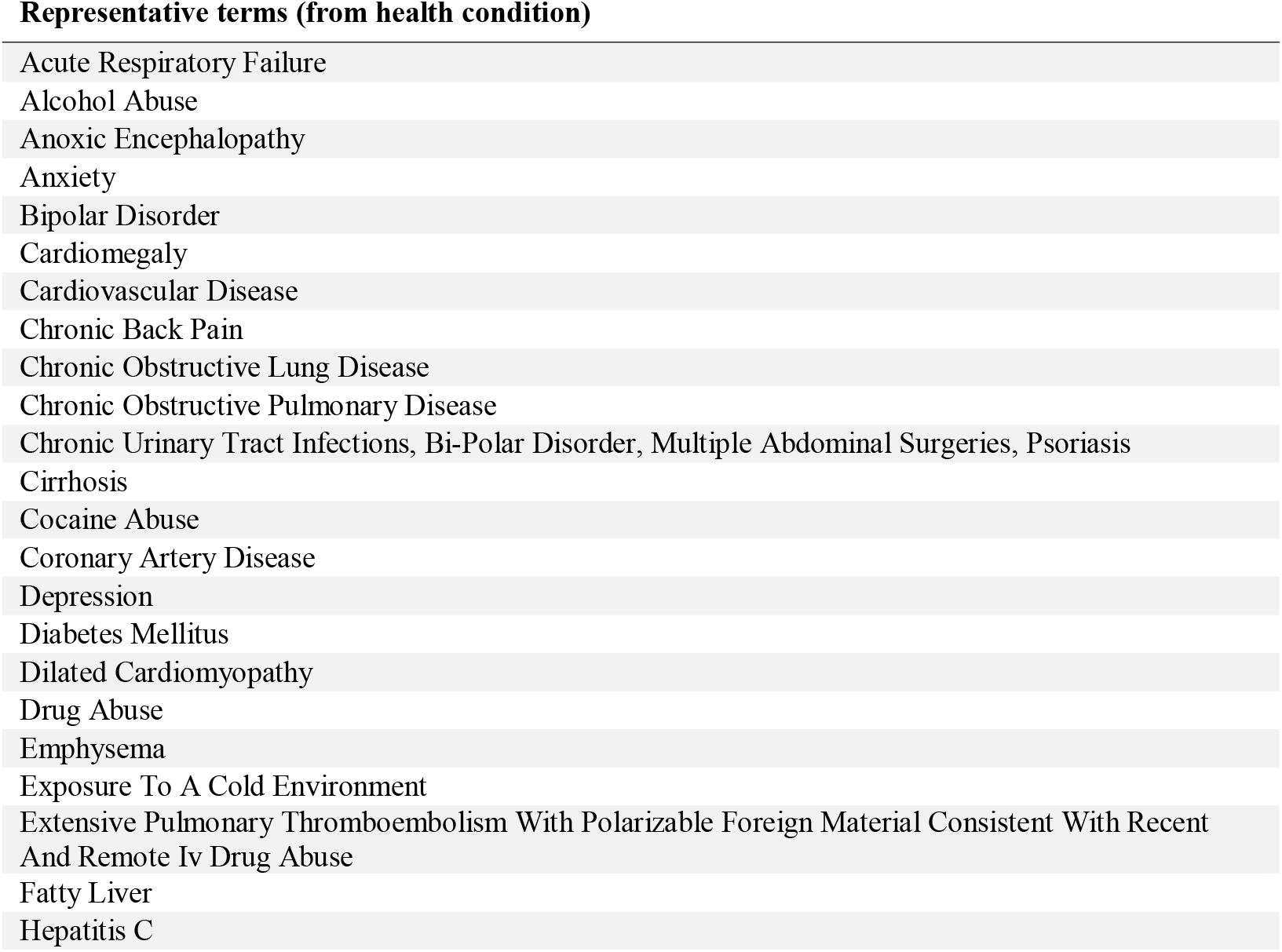
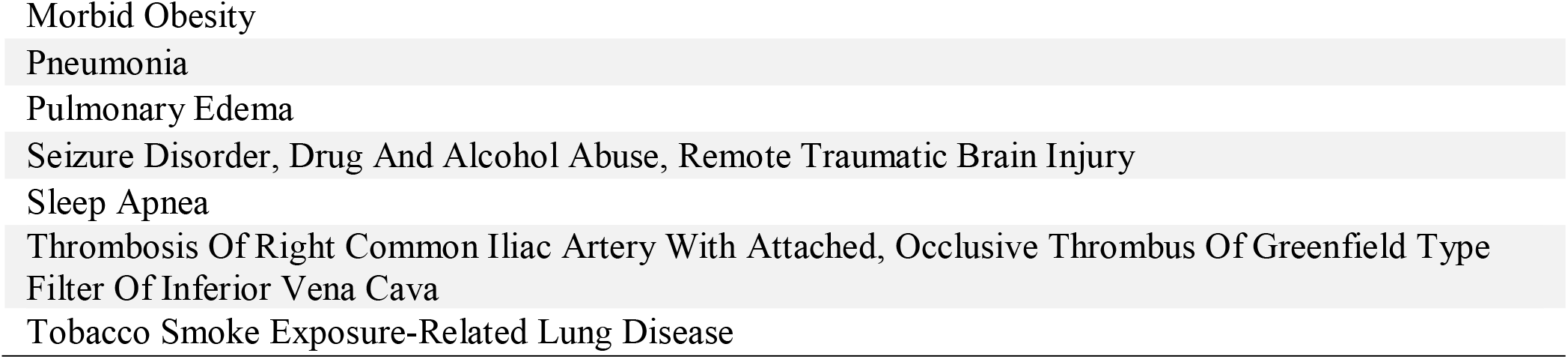
Results of clustering health conditions. We set the number of clusters to 70 through several empirical experiments. For each cluster, we selected the most frequent term, and then redundant clusters were manually removed by an expert. Through this post-processing, we could obtain 31 representative terms which indicates a given decedent’s health condition.

Frequent itemset mining was performed after setting the minimum support count to 200. From the results, the itemsets which had more than two items including one or more health conditions were selected. Table 4 shows the top 10 results based on frequency for this analysis. We revealed that cardiovascular disease was the most prevalent concomitant health condition in decedents by opioid poisoning. Cardiovascular disease frequently occurred with death due to hypertensive heart disease without heart failure. Drug abuse frequently occurred with death due to mental and behavioral disorders because of multiple drug use. Additionally, mental health conditions, such as anxiety, were frequently associated with opioid poisoning mortality.

**Table 4.** List of top 10 most frequent results which have more than two items including one or more health conditions in frequent itemset. We could identify that cardiovascular disease was the most prevalent concomitant health condition in decedents.

| Frequent Itemset | Frequency | Count | Description for cause of death | Description for health condition |
| --- | --- | --- | --- | --- |
| X42, clu_4 | 0.168 | 483 | Accidental poisoning by and exposure to narcotics and psychodysleptics [hallucinogens], not elsewhere classified | Cardiovascular disease |
| I119, clu_4 | 0.149 | 429 | Hypertensive heart disease without (congestive) heart failure | Cardiovascular disease |
| T509, clu_4 | 0.139 | 400 | Poisoning: Other and unspecified drugs, medicaments and biological substances | Cardiovascular disease |
| T402, clu_4 | 0.113 | 325 | Poisoning: Other opioids | Cardiovascular disease |
| T509, clu_23 | 0.112 | 324 | Poisoning: Other and unspecified drugs, medicaments and biological substances | Anxiety |
| I250, clu_4 | 0.110 | 319 | Atherosclerotic cardiovascular disease, so described | Cardiovascular disease |
| F191, clu_1 | 0.109 | 314 | Mental and behavioral disorders due to multiple drug use and use of other psychoactive substances: harmful use | Drug abuse |
| T401, clu_4 | 0.103 | 297 | Poisoning: Heroin | Cardiovascular disease |
| X42, clu_1 | 0.102 | 295 | Accidental poisoning by and exposure to narcotics and psychodysleptics [hallucinogens], not elsewhere classified | Drug abuse |
| I119, X42, clu_4 | 0.098 | 283 | Hypertensive heart disease without (congestive) heart failure<br>Accidental poisoning by and exposure to narcotics and psychodysleptics [hallucinogens], not elsewhere classified | Cardiovascular disease |

Table 5 shows the results of association rule mining. We identified that drug abuse, anxiety and cardiovascular disease were significantly and positively correlated with opioid poisoning mortality.

**Table 5.** List of the selected association rules which contain health condition in itemset *A* and cause of death in itemset *B* ordered by Lift value (≥1).

| Rule: $A \rightarrow B$ | | Confidence<br>($> 0.5$) | Lift (correlation for<br>dependency or<br>independency between<br>A and B) | Detailed description for rules |
| --- | --- | --- | --- | --- |
| itemset<br>A | itemset<br>B |  |  |  |
| clu_6 | I517 | 0.737 | 7.161 | Cardiomegaly → Cardiomegaly |
| clu_4 | I119 | 0.542 | 3.396 | Cardiovascular Disease → Hypertensive heart disease without (congestive) heart failure |
| clu_1 | F191 | 0.631 | 2.754 | Drug abuse → Mental and behavioral disorders due to multiple drug use and use of other psychoactive substances: harmful use |
| clu_23 | T402 | 0.504 | 1.364 | Anxiety → Poisoning: Other opioids |
| clu_1 | T401 | 0.502 | 1.322 | Drug abuse → Poisoning: Heroin |
| clu_23 | T509 | 0.582 | 1.117 | Anxiety → Poisoning: Other and unspecified drugs, medicaments and biological substances |
| clu_1 | T509 | 0.558 | 1.072 | Drug abuse → Poisoning: Other and unspecified drugs, medicaments and biological substances |
| clu_4 | X42 | 0.612 | 1.053 | Cardiovascular disease → Accidental poisoning by and exposure to narcotics and psychodysleptics [hallucinogens], not elsewhere classified |
| clu_1 | X42 | 0.592 | 1.020 | Drug abuse → Accidental poisoning by and exposure to narcotics and psychodysleptics [hallucinogens], not elsewhere classified |

## DISCUSSION

The opioid epidemic is one of the most pressing issues affecting Ohio, and news reports of overdose deaths constantly remind us of this. In August 2019, six people died of suspected fentanyl overdose within 24 hours, bringing the opioid-overdose death count to 10 for a three-day period.[23] All 10 deaths occurred within Cuyahoga County, Ohio, which encompasses Cleveland. This horrific event reminds us as researchers of the crucial need for research to prevent more deaths due to opioid overdose. Thus, with the accessibility to opioid abuse and overdose data, it is imperative to apply computational analysis to understanding the opioid epidemic.

To the best of our knowledge, there are currently no large-scale data driven approaches covering comprehensive analysis including profiling, spatial pattern mining, and association rule mining with statewide mortality records for all opioid-related deaths. Our research provides a comprehensive analysis in these areas, showing that various data mining algorithms such as text mining, spatial clustering and frequent pattern mining can be used to predict opioid overdose. The opioid epidemic is not only an important issue for research in the public health field, but also in the computer science field.

The first approach of our three-pronged analysis utilized demographic profiling and relative proportions to find disparities in opioid abuse among population groups. We found that people aged 25-44 and 45-54 died due to opioids more frequently than any other age group, and that whites and males also had higher rates of death due to opioid overdose. Our second approach used spatial autocorrelation to map geographical patterns of opioid abuse, and we found that opioid-related deaths have become more spatially concentrated over time. In particular, Cleveland, Columbus and Cincinnati had the highest concentration of deaths. Finally, our third approach focused on identifying health conditions that co-occurred with opioid-related death. We found that drug abuse, anxiety and cardiovascular disease were significantly and positively correlated with death due to opioid overdose.

The increasing spatial concentration of opioid overdose deaths that we observed is an interesting finding. From 1999-2004, the rate of opioid-related death rates in rural areas exceeded that in urban areas,[24] but recent data from the Centers for Disease Control and Prevention (CDC) shows an increase of drug overdose deaths in urban areas compared to that of rural areas in 2017.[25] Our results are consistent with this national pattern, which may suggest that Ohio undergoes similar changes in the spatial distribution of opioid abuse, and/or that our spatiotemporal model can be used as a tool to predict patterns of opioid use in consecutive years. Predictive geographical models would be essential in targeting prevention strategies for specific regions, as well as allocating resources to areas most in need.

Our comorbidity findings are consistent with existing studies that have identified anxiety, mental illness, and drug use to be associated with opioid abuse.[7,8,16-18] However, to our knowledge, other studies have not found an association between opioid abuse and cardiovascular disease. This finding may be due to the limited information from death records; only 22% contained information in the health condition field. In future studies, we plan to collect more robust data by combining vital records from ODH data and patient Electronic Health Record (EHR) data at the Cleveland Clinic. We recommend that similar studies be performed with data from hospitals in Columbus and Cincinnati to understand larger patterns. Such analyses can be used to develop a predictive model to stratify patient risk based on previously identified predictors of opioid misuse, which will aid caregivers in prescription decision-making.

Furthermore, we suggest specific application of spatial pattern mining to be used to identify areas of opioid abuse real-time, rather than solely in a retrospective fashion. A real-time predictive model would be indispensable for law enforcement agents and emergency health service providers to predict and prevent opioid abuse.

## CONCLUSION

As data related to opioid abuse and overdose have recently become available, large-scale, computational approaches to analysis are an imperative tool to combat the opioid epidemic. In this study, we utilized seven years of Ohio vital statistics data to reveal the race, age group, and regional differences of opioid poisoning. Additionally, we identified several prevalent health conditions that are frequently associated with opioid-related deaths. Although the data we utilized was limited to Ohio and was retrospective in nature, our findings help to understand the patterns and factors associated with opioid-related deaths, which could be useful in the development of predictive models. We expect that our findings provide essential knowledge to identify predictors of opioid misuse and predict the changing geographic pattern of opioid overdoses, enabling government officials and caregivers to improve their strategies for prevention and treatment of opioid abuse and to allocate resources most efficiently.

## Data Availability

Data was obtained from the publicly available U.S. Census datasets and American Community Survey.
Mortality data was obtained from the Ohio Department of Health. Access and use was approved by the ODH IRB.

https://www.census.gov/programs-surveys/acs/data.html

## ACKNOWLEDGMENTS

The authors would like to acknowledge Jesse Schold and Megan Snair of the Center for Populations Health Research of the Lerner Research Institute for their collaboration, Nabhonil Kar for his assistance on the manuscript, and the Ohio Department of Health for access to vital statistics data.

## Contributorship Statement

Conception or design of the work: GA, THH

Data analysis and interpretation: CP, SC, JRC, RL, TC, AS

Drafting the article: CP, AS, JRC, GA, THH

Critical revision of the article: CP, AS, JRC, GA, THH

Final approval of the version to be published: GA, THH

